# Using multivariate models to examine the impact of COVID-19 pandemic and gender differences on health and health care

**DOI:** 10.1101/2021.09.02.21263055

**Authors:** Jiancheng Ye, Zhimei Ren

## Abstract

**Objective:** To examine the effect of the COVID-19 pandemic, the effect of sex, and the joint effect of sex and the COVID-19 pandemic in relation to health communication, physical activity, mental health, and behavioral health.

**Methods:** We drew data from the National Cancer Institute’s 2020 Health Information National Trends Survey (HINTS). We described and compared the characteristics of social determinants of health, physical activity, mental health, alcohol use, patterns of social networking service use, and health information data sharing. Analyses were weighted to provide nationally representative estimates. Multivariate models (multiple linear regression, multiple logistic regression, and multinomial logistic model) were used to assess the sole and joint effect of sex and pandemic. In addition, we applied the Bonferroni correction to adjust p-values to decrease the risks of type I errors when making multiple statistical tests.

**Results:** Women are more likely to use mobile health and health communication technologies. The effect of sex after the COVID-19 pandemic is significant on mental health, and women are more possible to have depression or anxiety disorders. The effect of sex is also significant before and after the pandemic regarding seeking health or medical information. Women have a smaller quantity and intensity of physical activity, which has a negative effect on health.

**Conclusion:** Gender differences exist regardless of the COVID-19 pandemic and the pandemic amplifies the differences in some health and health care domains. Intersectional gender analyses are integral to addressing issues that arise and mitigating the exacerbation of inequities. Responses to the pandemic should consider diverse perspectives, including sex and gender.

## INTRODUCTION

The coronavirus disease 2019 (COVID-19) pandemic has uprooted conventional health care delivery for various care, such as primary care and mental health care, requiring health systems to rapidly adopt new capabilities.[1] With mandated social distancing policies in place during the COVID-19 pandemic, health care providers have been forced to get closely acquainted with virtual health. Virtual and remote care offers benefits such as accessibility. Additional benefits for both patients and providers can come from effectively leveraging patient-generated health data (PGHD) during these visits.[2] However, evidence shows mixed results regarding whether the widespread transition to telehealth during the pandemic is creating additional fractures in society by disproportionately harming health equity.[3]

Gender is one of the social determinants of health, unique from but entangled with sex differences and an axis along which the COVID-19 pandemic is widening health disparities. Regardless of the pandemic, women on average have reported more challenging physical and mental unhealthy status than men.[4] For example, evidence shows that more men than women are dying, potentially due to sex-based immunological or gendered differences, such as the prevalence of smoking.[5] The pandemic has differential impacts on women and men from the risks of exposure and biological susceptibility to infection, social and economic implications, and individuals’ experiences. These impacts are likely to vary according to biological and gender characteristics and their interaction with other social determinants.[6]

Recognizing the extent to which the pandemic affects women and men differently is a fundamental step to understanding the effects of the global public health emergency on different individuals and communities, and to creating effective interventions and equitable policies. The COVID-19 pandemic has further exacerbated inequities in health care, including patients’ access to necessary care and relevant health information. [7] There are also sex differences in health information technology (HIT) acceptability, physical activity, and mental health.[8] The digital divide has shown lower rates of technology and broadband adoption among lower socioeconomic statuses.[9]

Although a lot of work has studied the impact of COVID-19 or gender disparities,[10, 11] there is a lack of research on the sole and joint effect of COVID-19 or gender differences and their specific impact on health and health care during the pandemic. The aim of this study is to examine the effect of COVID-19 pandemic, the effect of sex, and the joint effect of sex and COVID-19 pandemic in relation to health communication, physical activity, mental health, and behavioral health.

## METHODS

### Study design

Data for this study were drawn from the National Cancer Institute’s 2020 Health Information National Trends Survey (HINTS). HINTS is a nationally representative survey administered every year by the National Cancer Institute, which provides a comprehensive assessment of the American public’s current access to and use of health information.[12] The target population of HINTS is civilian, non-institutionalized adults aged 18 or older living in the United States. In this study, we aim to examine the effect of the COVID-19 pandemic, the effect of sex, and the joint effect of sex and COVID-19 pandemic in relation to health communication, physical activity, and mental health, and behavioral health.

### Study participants

Data used in this study were from the third round of data collection for HINTS 5 (Cycle 4), which began in February 2020 and concluded in June 2020. A binary variable indicating whether the participants returned their survey before or after the COVID-19 pandemic was provided. This variable facilitates the examination of responses before and after COVID-19 in the United States. The final HINTS 5, Cycle 4 (2020) sample consists of 3,865 respondents.

### Model

To understand the effect of the pandemic, the effect of sex, and the joint effect of sex and pandemic, we characterize the response of interest via a multivariate model.[13] Specifically, If *Y* is continuous, we assume it to be generated from the following linear model:

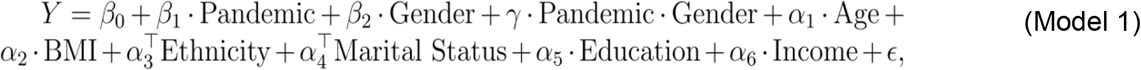

The *Pandemic* is a dichotomous variable indicating whether the response was obtained before or after the declaration of the pandemic; *Gender* is a dichotomous variable with 0 referring to the male and 1 the female; *ϵ* is a zero-mean normal random variable. Sociodemographic variables are incorporated in the model to account for possible confounding factors, where *Age* and *Income* are treated as continuous variables; *BMI, Ethnicity, Education Marital Status* are categorical variables. In model 1 *β*_1_ is the difference between the mean of the response after and before the pandemic for the males (adjusting the demographic covariates); *β*_1_ + *γ* is the same difference but in the female population. *β*_2_ represents the difference of the mean of the response between females and males (adjusting the demographic covariates) before the pandemic, *β*_2_ + *γ* and is the difference after the pandemic. The coefficient *γ* stands for the change of the gender difference before and after the pandemic, that is, (*β*_2_ + *γ*) − *β*_2_.

When the response *Y* is dichotomous (Y=0 is the reference), we build model 2, where we assume *Y* to be generated from a logistic regression model:

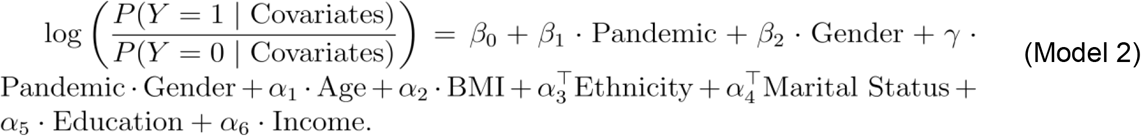

In model 2, the exp(*β*_1_) is the odds ratio (OR) of the male population after and before the pandemic adjusting the demographic covariates, and the exp(*β*_1_ + *γ*) is for the female population. The exp(*β*_2_) stands for the OR between females and males before the pandemic, exp(*β*_2_ + *γ*) and is the OR between females and males after the pandemic. The cxp(*γ*) is the ratio of the gender odds ratio after and before the pandemic, that is, exp(*β*_2_ + *γ*)/ exp(*β*_2_).

When the response *Y* is a categorical variable with more than two classes, we consider a multinomial logistic regression model.[14] Supposing the categories of the *Y* are denoted by the set= {1, 2, …, *K*}, we set the first category as the baseline, for each *k* = 2, …, *K* and build model 3:

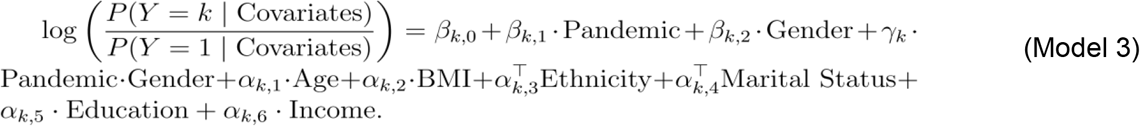

The interpretation of model 3 is the same as model 2 except that we are comparing category k to the reference category.

### Statistical Analysis

Statistical analyses were performed using R version 4.0.5 (R Foundation, Vienna, Austria). For continuous responses, ordinary least squares (OLS) estimators of the coefficients in the linear model are estimated with the lm function in R and reported with the p-values; for categorical responses, adjusted odds ratios (aORs) are estimated with the glm package in R, and reported with the p-values. The p-values for the sum of two coefficient estimators are obtained using the multcomp package in R, and all statistical testing was 2-tailed. In this study, 50 characteristics were categorized into four domains (health communication, physical activity, mental health, and behavioral health). Within each domain, there were five hypotheses to be tested: the effect of sex before the pandemic, the effect of sex after the pandemic, the effect of the pandemic in male population, the effect of the pandemic in female population, and the interacting effect of gender and pandemic.

Commonly, p=0.05 is used as the significance threshold for a single hypothesis testing. In this study, 50*5=250 hypotheses are tested simultaneously. Therefore, it is necessary to adjust for the multiplicity of the hypotheses, to avoid type I errors.[15] Assuming that all the 250 hypotheses are null, if we are still using the 0.05 threshold, then the probability of making at least a false discovery is the probability of at least one p-value exceeding 0.05. When the p-values are mutually independent, the value is 1-(0.95) ^ 250 ≈ 1.00, which means we almost surely make at least one false discovery. Meanwhile, the expected number of discoveries is 250*0.05=12.5, and these discoveries are all, by definition, false discoveries. To address this issue, we need to adjust the p-value threshold based on the number of hypotheses being tested. In particular, we apply the Bonferroni correction [16] to adjust p-values to decrease the risks of type I errors, which proceeds as the following steps: when testing m hypotheses simultaneously, a p-value threshold of 0.05/m is utilized. With the adjusted threshold, the *m*_0_ denotes the number of null hypotheses, and the probability of making at least one false discovery can be bounded as:

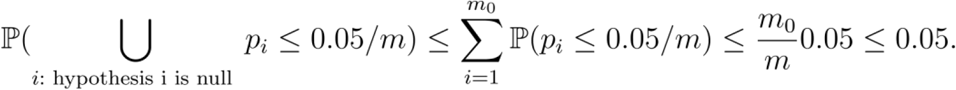

The above derivation illustrates that the probability of making at least one false discovery is no more than 0.05, which is desirable. As a result, a p-value < 0.05/250=0.0002 is designated as statistically significant in our statistical analyses. We indicated the significant variables in the results section and all the p-values for Table 2 - 4 and figure 1 are shown in the **supplemental tables**.

**Figure 1.**
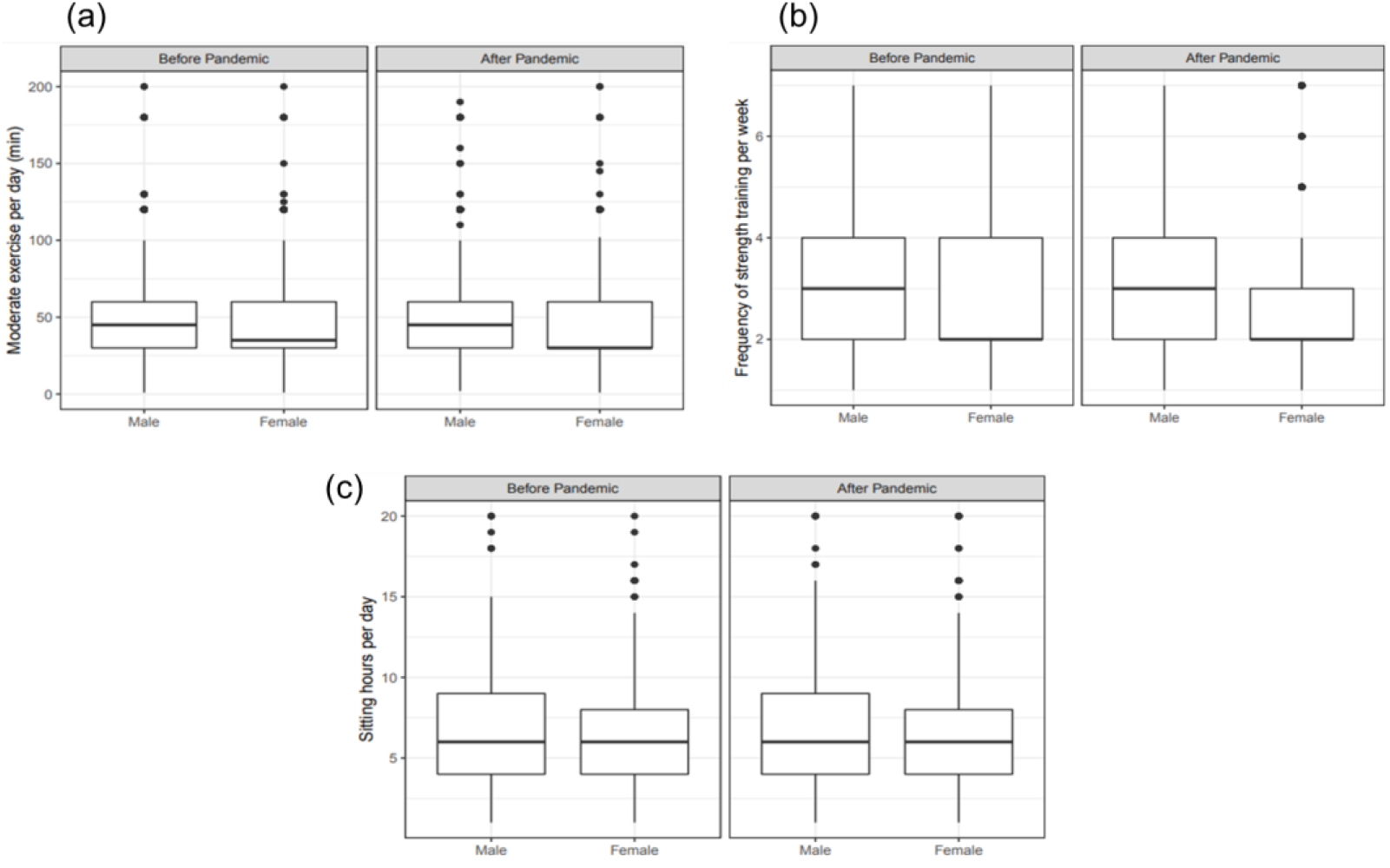
Multivariate modeling the effect of COVID-19 pandemic, the effect of sex, and the joint effect of sex and COVID-19 pandemic in relation to physical health

## RESULTS

Table 1 demonstrates the sociodemographic characteristics of the participants in this study. We present the weighted and unweighted prevalence of subcategories for each variable, and the weighted prevalence of the subcategories before and after the COVID-19 pandemic. There were more younger participants and white participants after the pandemic, but the other sociodemographics didn’t show significant differences.

**Table 1.** Unweighted and weighted prevalence estimates for sample sociodemographic, Health Information National Trends Survey (HINTS) 5 Cycle 4 (2020)

| Characteristics | Overall Unweighted, % | Overall Weighted, % | Before pandemic, Weighted, % | After pandemic, Weighted, % | P-Value |
| --- | --- | --- | --- | --- | --- |
| **Age, %** |  |  |  |  | <0.001 |
| 18–34 | 12.9 | 26.2 | 19.9 | 29.6 |  |
| 35–49 | 18.8 | 25.6 | 22.7 | 27.1 |  |
| 50–64 | 30.6 | 27.7 | 33.0 | 24.8 |  |
| 65+ | 37.6 | 20.5 | 24.4 | 18.4 |  |
| **Male, %** | 41.5 | 48.6 | 51.0 | 47.4 | 0.27 |
| **BMI, %** |  |  |  |  | 0.36 |
| Underweight | 1.7 | 1.3 | 1.4 | 1.2 |  |
| Normal weight | 31.6 | 34.1 | 33.0 | 34.6 |  |
| Overweight | 33.7 | 31.5 | 34.0 | 30.1 |  |
| Obesity | 33.1 | 33.2 | 31.6 | 34.1 |  |
| **Race/Ethnicity, %** |  |  |  |  | <0.001 |
| Non-Hispanic White | 61.3 | 63.4 | 70.1 | 59.8 |  |
| Non-Hispanic Black or African American | 13.7 | 11.0 | 8.1 | 12.6 |  |
| Hispanic | 16.9 | 16.9 | 13.0 | 19.1 |  |
| Non-Hispanic Asian | 4.7 | 5.3 | 4.2 | 5.8 |  |
| Non-Hispanic Other | 3.4 | 3.3 | 4.6 | 2.7 |  |
| **Marital status, %** |  |  |  |  | 0.17 |
| Married | 48.7 | 50.6 | 54.5 | 48.6 |  |
| Divorced | 16.1 | 8.3 | 8.2 | 8.4 |  |
| Widowed | 11.0 | 4.6 | 5.2 | 4.3 |  |
| Other† | 24.2 | 36.4 | 32.1 | 38.8 |  |
| **Education, %** |  |  |  |  | 0.15 |
| High school diploma or less | 33.2 | 39.2 | 38.2 | 39.8 |  |
| Some college | 21.9 | 30.3 | 28.4 | 31.3 |  |
| College graduate or higher | 44.9 | 30.5 | 33.4 | 28.9 |  |
| **Income, %** |  |  |  |  | 0.58 |
| Less than \$20,000 | 17.8 | 14.9 | 13.6 | 15.5 | |
| \$20,000–\$34,999 | 13.0 | 11.4 | 10.5 | 11.8 | |
| \$35,000–\$49,999 | 13.4 | 12.7 | 12.2 | 13.0 | |
| \$50,000–\$74,999 | 17.3 | 18.4 | 17.8 | 18.8 | |
| \$75,000 or more | 38.6 | 42.7 | 45.9 | 40.9 | |

†Other includes single (never been married), separated, living as married or living with a romantic partner.

Table 2 presents the estimates of the multivariate model in terms of the variables corresponding to the health communication. For the characteristic *made appointments with a health care provider online*, the effect of sex before the pandemic is not significant but becomes significant after the pandemic, where the aOR between the female and male is e^-0.36^=0.70 (the other aORs in the tables can be interpreted in the same way), which means the odds of making appointments with a health care provider online for males is 0.7 times that of females; the effect of the pandemic is not significant either before or after the pandemic, and the interacting effect of the pandemic and sex is not significant either. As for characteristic *looked for health or medical information for oneself*, the effect of sex is significant before the pandemic, with an aOR=e^-0.65^=0.52, which means the odds of looking for health or medical information for males is 0.52 times of the female; the effect of sex is also significant after the pandemic, with an aOR between the female and male population is e^-0.55^= 0.58. Similarly, we see a significant effect of sex both before and after the pandemic in characteristics including *looked up medical test results, shared health information on social networking sites, participated in an online forum or support group, visited social networking sites, used smartphone to track progress on a health-related goal* and *used an electronic wearable device to monitor or track health or activity*. We also see a significant effect of sex after the pandemic with regard to the characteristic *have “apps” related to health and wellness*. We see gender differences in using mobile health and social networking services, and females are more likely to use these tools.

**Table 2.**
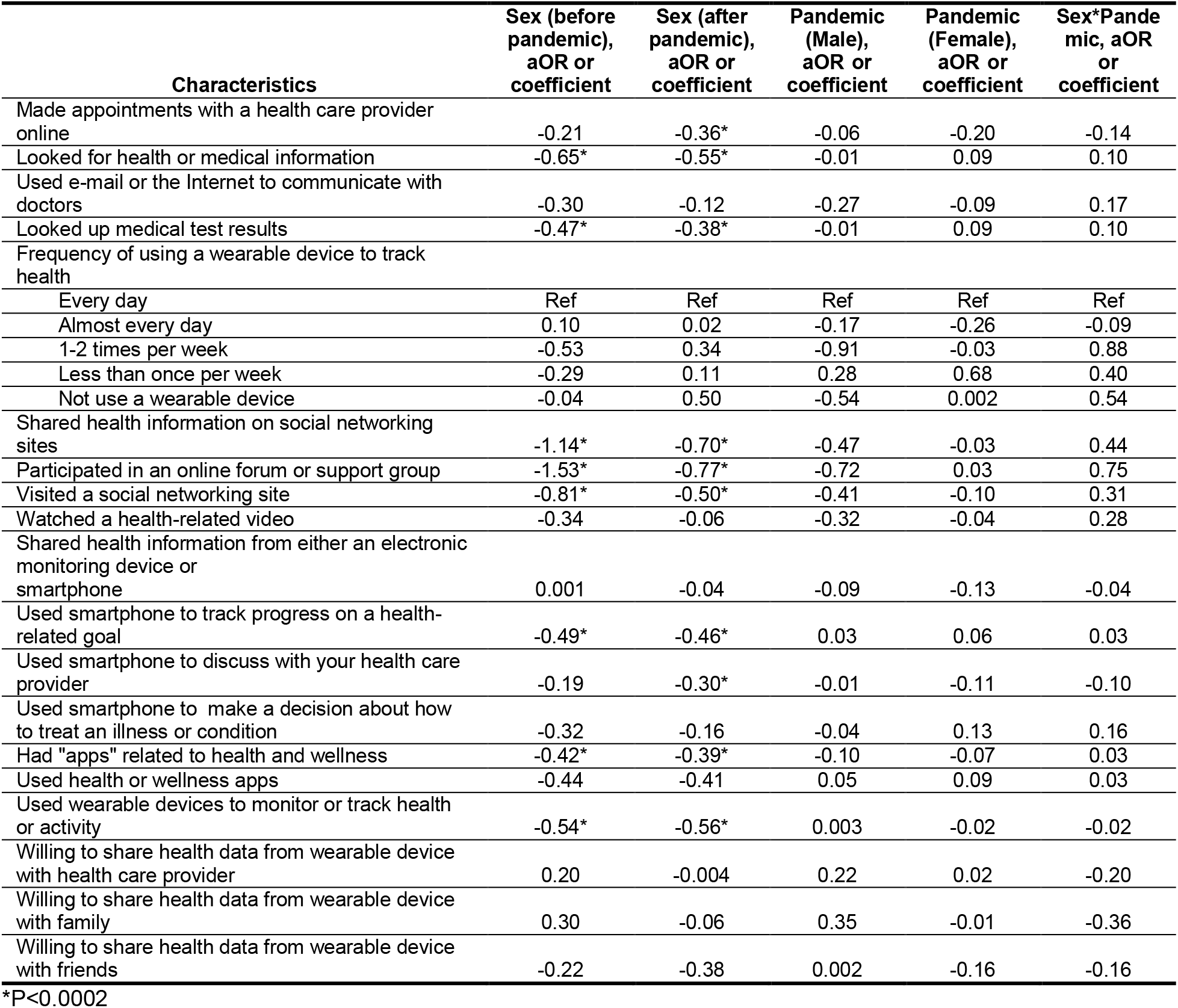
Multivariate modeling the effect of COVID-19 pandemic, the effect of sex, and the joint effect of sex and COVID-19 pandemic in relation to health communication

Table 3 presents the model estimates for the characteristics corresponding to mental health. For *Depression or Anxiety*, the effect of sex is significant before and after the pandemic, with estimated effects being -0.09 and -0.13, respectively. This means the difference of the levels of having depression or anxiety disorder in females and males are -0.09 and -0.31 before and after the pandemic respectively. For *Feeling nervous, anxious, or on edge*, the effect of sex is significant before and after the pandemic, with estimated coefficient -0.15 and -0.19 respectively, suggesting that females are more likely to feel nervous or anxious. The effect of sex demonstrates a significant effect for *not being able to stop or control worrying* after the pandemic, with an expected difference -0.19. Sex also shows a significant effect for characteristics like *When I feel threatened or anxious, I find myself thinking about my strengths* before the pandemic, with an expected difference -0.17. The characteristic *Most important value* is categorical with more than two classes. As introduced in the Methods section, we set *Making my own decisions* as the reference, and compare other values to it. The effect of sex is significant after the pandemic for the characteristics *being happy, having a deep connection to my religion and assuring my family is safe and secure* (when compared with the reference), and the aORs are 3.53 and 1.93 respectively. Overall, females are more possible to have mental health issues

**Table 3.** Multivariate modeling the effect of COVID-19 pandemic, the effect of sex, and the joint effect of sex and COVID-19 pandemic in relation to mental health

| Characteristics | Sex (before pandemic), aOR or coefficient | Sex (after pandemic), aOR or coefficient | Pandemic (Male), aOR or coefficient | Pandemic (Female), aOR or coefficient | Sex*Pandemic, aOR or coefficient |
| --- | --- | --- | --- | --- | --- |
| General health | -0.003 | 0.02 | 0.02 | 0.04 | 0.02 |
| Confident about ability to take good care of health | -0.04 | -0.06 | 0.05 | 0.03 | -0.02 |
| Depression or anxiety disorder | -0.09* | -0.13* | 0.01 | -0.02 | -0.03 |
| Little interest or pleasure in doing things | -0.01 | -0.01 | 0.003 | -0.001 | -0.004 |
| Feeling down, depressed, or hopeless | -0.11 | -0.06 | -0.03 | 0.01 | 0.05 |
| Feeling nervous, anxious, or on edge | -0.15* | -0.19* | -0.02 | -0.06 | -0.04 |
| Not being able to stop or control worrying | -0.23 | -0.19* | -0.02 | 0.01 | 0.03 |
| When I feel threatened or anxious I find myself thinking about my values | -0.14 | -0.08 | -0.10 | -0.04 | 0.06 |
| When I feel threatened or anxious I find myself thinking about my strengths | -0.17* | -0.06 | -0.14 | -0.03 | 0.11 |
| Most important value |  |  |  |  |  |
| Making my own decisions | Ref | Ref | Ref | Ref | Ref |
| Being happy | 0.01 | 0.54 | -0.44 | 0.09 | 0.53 |
| Helping people | 0.40 | 0.59 | -0.01 | 0.17 | 0.19 |
| Being loyal to family and friends | 0.23 | -0.19 | -0.13 | -0.56 | -0.43 |
| Having a deep connection to my religion | 0.46 | 1.26* | -0.45 | 0.35 | 0.80 |
| Keeping myself in good health | 0.08 | 0.40 | -0.17 | 0.15 | 0.32 |
| Assuring my family is safe and secure | 0.03 | 0.66* | -0.21 | 0.42 | 0.63 |

\*P<0.0002

Table 4 presents the model estimates for variables corresponding to behavioral health. In this table, *Number of days drinking alcohol* and *Average drinks per day* are treated as continuous variables, and *Frequency of alcohol use per month* is treated as a categorical variable. For the number of days drinking alcohol, the effect of sex is significant before and after the pandemic, with estimated coefficients -0.59 and -0.39, respectively. Similarly, there is a significant effect of sex for *average drinks per day*, before and after the pandemic. The characteristic *Frequency of alcohol use per month* is a categorical variable with 5 classes. We set *Never* as the reference and compare other classes to the reference, and there are no significant effects either from sex or the pandemic.

**Table 4.** Multivariate modeling the effect of COVID-19 pandemic, the effect of sex, and the joint effect of sex and COVID-19 pandemic in relation to behavioral health

| Characteristics | Sex (before pandemic), aOR or coefficient | Sex (after pandemic), aOR or coefficient | Pandemic (Male), aOR or coefficient | Pandemic (Female), aOR or coefficient | Sex*Pandemic, aOR or coefficient |
| --- | --- | --- | --- | --- | --- |
| # of days drinking alcohol | -0.59* | -0.39* | -0.14 | 0.06 | 0.20 |
| Average drinks per day | -0.64* | -0.67* | -0.18 | -0.20 | -0.03 |
| Frequency of alcohol use per month |  |  |  |  |  |
| Never | Ref | Ref | Ref | Ref | Ref |
| 1-2 | -0.35 | -0.24 | 0.03 | 0.14 | 0.11 |
| 3-5 | -0.20 | -0.62 | 0.37 | -0.05 | -0.42 |
| 6-10 | -0.78 | -0.39 | -0.39 | -0.01 | 0.39 |
| 11+ | -0.83 | -0.18 | -0.21 | 0.44 | 0.65 |

\*P<0.0002

We also examined the effects of sex and pandemic on physical activity. Figure 1 shows the three measurements of physical activities in the male and female population, before and after the pandemic.

Figure 1 (a) compares the minutes of moderate exercise per day, where outliers (minutes >= 200) are removed. The effect of sex is significant for moderate exercise per day after the pandemic. Figure 1 (b) compares the frequency of strength training per week, and we see that males tend to have a higher frequency of strength training regardless of the pandemic. Figure 1 (c) compares the sitting time per day, showing that males sit slightly longer than females regardless of the pandemic.

## DISCUSSION

Gender disparities in health care could be defined as the dominance of one gender over the other in particular contexts, such as among primary social roles or occupations. Stereotypes appear when the less dominant genders are always underrepresented.[17] The biological, behavioral, social, and systemic factors underlying the differences in how women and men may experience COVID-19 and its consequences cannot be oversimplified.[18] Responses to the pandemic should consider diverse perspectives, including sex and gender. Individuals seek information for informed decision-making, and they consult a variety of information sources nowadays. Although we do not see a significant effect of the interaction term for all the characteristics, suggesting that the pandemic has not statistically affected the two genders differently. However, the results show that the pandemic amplifies some existing gender differences in health information seeking, communication, sharing, physical activity, and mental health. To avoid perpetuating gender and health inequities in responses to public health emergencies such as COVID-19 is not perpetuate gender and health inequities, it is important that gender norms, roles, and relations that influence women’s and men’s differential health communication behaviors are considered and addressed.[19] Early evidence is mixed regarding whether the widespread transition to telehealth during the pandemic is creating additional fractures in society by disproportionately influencing health equity. [3] Our results show that implementation of remote health care or using mobile health needs to be grounded in strong gender analysis and ensure meaningful participation of affected groups, including women.

The pandemic induces a slight decrease in the quantity and intensity of physical activity in both genders, as well as an increase in alcohol drinking, which has a negative effect on health.[20] We found that women have a lower level of physical activity than men, and this gap has slightly increased during the pandemic. The effect of sex is significant before and after the pandemic regarding alcohol assumption, and men drink more than women. However, it is necessary to highlight that the pandemic has a greater negative impact on women.

In terms of mental health, the COVID-19 pandemic can be experienced in many different ways, including feelings of depression, fear, panic, and anxiety. Stress-related disorders and the long-term consequences of COVID-19 on health outcomes highlight another important effect of sex and gender. COVID-19 acts as a potent stressor, with millions of individuals experiencing fear and social isolation over a prolonged period.[10] In this study, we found that women are more likely to have increased vulnerability to and severity of stress-related psychiatric disorders than men. These observations may be helpful to developing and implementing prevention and treatment interventions that are able to address the acute and long-term effects of the pandemic on the health and well-being of both male and female populations.

### LIMITATION

While the mailing and the reminder postcards were sent out on schedule and without any issues, the World Health Organization’s announcement on March 11 of the COVID-19 pandemic impacted the rest of the field period in HINTS 5 Cycle 4. Given the limitations of the dataset, we did not have information about when the participants filled out the survey, which may have impacted the results. Because the survey was cross-sectional, we could not examine causality among variables. Despite these limitations, this study provides nationally representative estimates and contributes to a better understanding of the effects of the gender differences and COVID-19 pandemic on common characteristics of health and health care.

## CONCLUSION

The COVID-19 pandemic has devastated marginalized communities, exposing the deep inequities of the health care system. This study used multivariate models to understand the effect of COVID-19 pandemic, the effect of sex, and the joint effect of sex and COVID-19 pandemic in relation to health communication, physical activity, mental health, and behavioral health. The findings of this study demonstrate that the COVID-19 pandemic amplifies existing gender disparities in some health and health care domains. Intersectional gender analyses are integral to addressing issues that arise and mitigating inequities. Responses to the pandemic should consider diverse perspectives, including sex and gender. Decisions that are informed by accurate data and include sex and gender perspectives are essential.

## Data Availability

All data referred to in the manuscript are available on: https://www.cancer.gov/, and the relevant code and analyses are available at: https://github.com/zhimeir/hints_analysis

## AUTHOR CONTRIBUTIONS

J.Y. conceived and designed the study, and contributed to the analyses. Z.R. contributed to the analyses. J.Y. and Z.R. contributed to the interpretation of the results, drafting, and revision of the manuscript. All the authors read and approved the final version of the manuscript.

