## Supplemental Tables for "Using multivariate models to examine the impact of COVID-19 pandemic and gender differences on health and health care"

**Table 1.** Multivariate modeling the effect of COVID-19 pandemic, the effect of sex, and the joint effect of sex and COVID-19 pandemic in relation to health communication

| Characteristics | Sex (before pandemic) | P-Value | Sex (after pandemic) | P-Value | Pandemic (Male) | P-Value | Pandemic (Female) | P-Value | Sex*Pandemic | P-Value |
|---|---|---|---|---|---|---|---|---|---|---|
| Made appointments with a health care provider online | -0.21 | 0.06135 | -0.36* | 0.00008 | -0.06 | 0.59721 | -0.20 | 0.03412 | -0.14 | 0.32426 |
| Looked for health or medical information | -0.65* | <0.00001 | -0.55* | <0.00001 | -0.01 | 0.95950 | 0.09 | 0.42727 | 0.10 | 0.55909 |
| Used e-mail or the Internet to communicate with doctors | -0.30 | 0.01115 | -0.12 | 0.17803 | -0.27 | 0.01791 | -0.09 | 0.34181 | 0.17 | 0.23926 |
| Looked up medical test results | -0.47* | 0.00006 | -0.38* | 0.00005 | -0.01 | 0.94491 | 0.09 | 0.36258 | 0.10 | 0.51833 |
| Frequency of using a wearable device to track health | | | | | | | | | | |
| Every day | Ref | Ref | Ref | Ref | Ref | Ref | Ref | Ref | Ref | Ref |
| Almost every day | 0.10 | 0.70690 | 0.02 | 0.93916 | -0.17 | 0.55238 | -0.26 | 0.20904 | -0.09 | 0.80127 |
| 1-2 times per week | -0.53 | 0.14888 | 0.34 | 0.30908 | -0.91 | 0.02377 | -0.03 | 0.92056 | 0.88 | 0.07811 |
| Less than once per week | -0.29 | 0.56776 | 0.11 | 0.73102 | 0.28 | 0.57159 | 0.68 | 0.05812 | 0.40 | 0.50286 |
| Not use a wearable device | -0.04 | 0.92075 | 0.50 | 0.10873 | -0.54 | 0.17108 | 0.002 | 0.99283 | 0.54 | 0.25518 |
| Shared health information on social networking sites | -1.14* | <0.00001 | -0.70* | <0.00001 | -0.47 | 0.02378 | -0.03 | 0.80334 | 0.44 | 0.07042 |
| Participated in an online forum or support group | -1.53* | <0.00001 | -0.77* | <0.00001 | -0.72 | 0.01109 | 0.03 | 0.83469 | 0.75 | 0.01841 |
| Visited a social networking site | -0.81* | <0.00001 | -0.50* | <0.00001 | -0.41 | 0.00047 | -0.10 | 0.34676 | 0.31 | 0.05569 |
| Watched a health-related video | -0.34 | 0.00553 | -0.06 | 0.53128 | -0.32 | 0.00560 | -0.04 | 0.66834 | 0.28 | 0.06364 |
| Shared health information from either an electronic monitoring device or smartphone | 0.001 | 0.99753 | -0.04 | 0.75350 | -0.09 | 0.54572 | -0.13 | 0.31021 | -0.04 | 0.84550 |
| Used smartphone to track progress on a health-related goal | -0.49* | 0.00012 | -0.46* | <0.00001 | 0.03 | 0.82181 | 0.06 | 0.58317 | 0.03 | 0.85772 |
| Used smartphone to discuss with your health care provider | -0.19 | 0.12214 | -0.30* | 0.00245 | -0.01 | 0.94105 | -0.11 | 0.26701 | -0.10 | 0.51356 |
| Used smartphone to make a decision about how to treat an illness or condition | -0.32 | 0.00936 | -0.16 | 0.09888 | -0.04 | 0.75389 | 0.13 | 0.20886 | 0.16 | 0.29528 |
| Had "apps" related to health and wellness | -0.42* | 0.00077 | -0.39* | 0.00007 | -0.10 | 0.41778 | -0.07 | 0.50636 | 0.03 | 0.85341 |
| Used health or wellness apps | -0.44 | 0.08267 | -0.41 | 0.03717 | 0.05 | 0.82551 | 0.09 | 0.68366 | 0.03 | 0.91551 |
| Used wearable devices to monitor or track health or activity | -0.54* | 0.00004 | -0.56* | <0.00001 | 0.003 | 0.98075 | -0.02 | 0.87789 | -0.02 | 0.90893 |
| Willing to share health data from wearable device with health care provider | 0.20 | 0.52385 | -0.004 | 0.98636 | 0.22 | 0.48429 | 0.02 | 0.94173 | -0.20 | 0.59670 |
| Willing to share health data from wearable device with family | 0.30 | 0.26287 | -0.06 | 0.77623 | 0.35 | 0.20028 | -0.01 | 0.96621 | -0.36 | 0.28039 |
| Willing to share health data from wearable device with friends | -0.22 | 0.32497 | -0.38 | 0.02799 | 0.002 | 0.99287 | -0.16 | 0.33824 | -0.16 | 0.56181 |

**Table 2.** Multivariate modeling the effect of COVID-19 pandemic, the effect of sex, and the joint effect of sex and COVID-19 pandemic in relation to mental health

| Characteristics | Sex (before pandemic) | P-Value | Sex (after pandemic) | P-Value | Pandemic (Male) | P-Value | Pandemic (Female) | P-Value | Sex*Pandemic | P-Value |
|---|---|---|---|---|---|---|---|---|---|---|
| General health | -0.003 | 0.94967 | 0.02 | 0.62858 | 0.02 | 0.61393 | 0.04 | 0.26003 | 0.02 | 0.72463 |
| Confident about ability to take good care of health | -0.04 | 0.41543 | -0.06 | 0.10051 | 0.05 | 0.19954 | 0.03 | 0.35946 | -0.02 | 0.70453 |
| Depression or anxiety disorder | -0.09* | 0.00004 | -0.13* | <0.00001 | 0.01 | 0.54033 | -0.02 | 0.34730 | -0.03 | 0.27986 |
| Little interest or pleasure in doing things | -0.01 | 0.88212 | -0.01 | 0.76149 | 0.003 | 0.94076 | -0.001 | 0.98131 | -0.004 | 0.94266 |
| Feeling down, depressed, or hopeless | -0.11 | 0.00796 | -0.06 | 0.05494 | -0.03 | 0.38855 | 0.01 | 0.69876 | 0.05 | 0.36271 |
| Feeling nervous, anxious, or on edge | -0.15* | 0.00123 | -0.19* | <0.00001 | -0.02 | 0.69036 | -0.06 | 0.11737 | -0.04 | 0.47110 |
| Not being able to stop or control worrying | -0.23 | <0.00001 | -0.19* | <0.00001 | -0.02 | 0.60430 | 0.01 | 0.78751 | 0.03 | 0.56776 |
| When I feel threatened or anxious I find myself thinking about my values | -0.14 | 0.01940 | -0.08 | 0.09588 | -0.10 | 0.07951 | -0.04 | 0.42451 | 0.06 | 0.41668 |
| When I feel threatened or anxious I find myself thinking about my strengths | -0.17* | 0.00233 | -0.06 | 0.17163 | -0.14 | 0.00715 | -0.03 | 0.45288 | 0.11 | 0.11836 |
| Most important value | | | | | | | | | | |
| Making my own decisions | Ref | Ref | Ref | Ref | Ref | Ref | Ref | Ref | Ref | Ref |
| Being happy | 0.01 | 0.94432 | 0.54 | 0.00112 | -0.44 | 0.02492 | 0.09 | 0.60410 | 0.53 | 0.04455 |
| Helping people | 0.40 | 0.17058 | 0.59 | 0.00718 | -0.01 | 0.96746 | 0.17 | 0.44913 | 0.19 | 0.61007 |
| Being loyal to family and friends | 0.23 | 0.37263 | -0.19 | 0.42773 | -0.13 | 0.60071 | -0.56 | 0.02284 | -0.43 | 0.22848 |
| Having a deep connection to my religion | 0.46 | 0.05605 | 1.26* | <0.00001 | -0.45 | 0.06824 | 0.35 | 0.06734 | 0.80 | 0.01054 |
| Keeping myself in good health | 0.08 | 0.69958 | 0.40 | 0.01448 | -0.17 | 0.36470 | 0.15 | 0.39988 | 0.32 | 0.21342 |
| Assuring my family is safe and secure | 0.03 | 0.89270 | 0.66* | 0.00005 | -0.21 | 0.26452 | 0.42 | 0.01782 | 0.63 | 0.01429 |

**Table 3.** Multivariate modeling the effect of COVID-19 pandemic, the effect of sex, and the joint effect of sex and COVID-19 pandemic in relation to behavioral health

| Characteristics | Sex (before pandemic) | P-Value | Sex (after pandemic) | P-Value | Pandemic (Male) | P-Value | Pandemic (Female) | P-Value | Sex*Pandemic | P-Value |
|---|---|---|---|---|---|---|---|---|---|---|
| # of days drinking alcohol | -0.59* | <0.00001 | -0.39* | 0.00001 | -0.14 | 0.21418 | 0.06 | 0.49409 | 0.20 | 0.16376 |
| Average drinks per day | -0.64* | 0.00005 | -0.67* | <0.00001 | -0.18 | 0.22417 | -0.20 | 0.14943 | -0.03 | 0.89792 |
| Frequency of alcohol use per month | | | | | | | | | | |
| Never | Ref | Ref | Ref | Ref | Ref | Ref | Ref | Ref | Ref | Ref |
| 1-2 | -0.35 | 0.07993 | -0.24 | 0.12569 | 0.03 | 0.84864 | 0.14 | 0.41695 | 0.11 | 0.66492 |
| 3-5 | -0.20 | 0.51482 | -0.62 | 0.00817 | 0.37 | 0.16111 | -0.05 | 0.86591 | -0.42 | 0.26662 |
| 6-10 | -0.78 | 0.14316 | -0.39 | 0.37907 | -0.39 | 0.40603 | -0.01 | 0.99197 | 0.39 | 0.57383 |
| 11+ | -0.83 | 0.08145 | -0.18 | 0.62269 | -0.21 | 0.58762 | 0.44 | 0.32668 | 0.65 | 0.27026 |

**Table 4.** Multivariate modeling the effect of COVID-19 pandemic, the effect of sex, and the joint effect of sex and COVID-19 pandemic in relation to physical health

| Characteristics | Sex (before pandemic) | P-Value | Sex (after pandemic) | P-Value | Pandemic (Male) | P-Value | Pandemic (Female) | P-Value | Sex*Pandemic | P-Value |
|---|---|---|---|---|---|---|---|---|---|---|
| Moderate exercise per day, mins | -11.27 | 0.00090 | -14.11* | <0.00001 | 1.44 | 0.65544 | -1.39 | 0.62191 | -2.83 | 0.50740 |
| Sitting hours per day | -0.33 | 0.11003 | -0.33 | 0.03952 | 0.00 | 0.98245 | -0.01 | 0.95375 | -0.01 | 0.98290 |
| Frequency of strength training per week | -0.17 | 0.08889 | -0.24 | 0.00258 | 0.02 | 0.84615 | -0.05 | 0.55224 | -0.07 | 0.59141 |
